# HPV and HBV vaccine hesitancy, intention, and uptake in the era of social media and COVID-19: A review

**DOI:** 10.1101/2023.01.25.23285015

**Authors:** Emily K Vraga, Sonya S Brady, Chloe Gansen, Euna M Khan, Sarah L. Bennis, Madalyn Nones, Rongwei Tang, Jaideep Srivastava, Shalini Kulasingam

**Author notes:** Corresponding Author: Emily Vraga, PhD, 338 Murphy Hall, 206 Church Street SE University of Minnesota, Minneapolis, MN 55455.

## Abstract

Prior to the COVID-19 pandemic, the World Health Organization named vaccine hesitancy as one of the top 10 threats to global health. The impact of hesitancy on uptake of human papillomavirus (HPV) vaccines was of particular concern, given the markedly lower uptake compared to other adolescent vaccines in some countries, notably the United States. With the recent approval of COVID-19 vaccines coupled with the widespread use of social media, concerns regarding vaccine hesitancy have grown. However, the association between COVID-related vaccine hesitancy and cancer vaccines such as HPV is unclear. To examine the potential association, we performed two reviews using Ovid Medline and APA PsychInfo. Our aim was to answer two questions: (1) Is COVID-19 vaccine hesitancy, intention, or uptake associated with HPV or HBV vaccine hesitancy, intention, or uptake? and (2) Is exposure to COVID-19 vaccine misinformation on social media associated with HPV or HBV vaccine hesitancy, intention, or uptake? Our review identified few published empirical studies that addressed these questions. Our results highlight the urgent need for studies that can shift through the vast quantities of social media data to better understand the link between COVID-19 vaccine misinformation and disinformation and its impact on uptake of cancer vaccines.

---

The issue of vaccine hesitancy is not a new one. For as long as there have been vaccines, there have been skeptics who distrusted the efficacy and safety of vaccination and who have shared misinformation about their value – and such misinformation is a major driver of vaccine hesitancy (Enders, Uscinski, Klofstad, & Stoler, 2022; Lee, Sun, Jang, & Connelly, 2022). We define misinformation as any information that counters the current best evidence and expert consensus on the topic (Vraga & Bode, 2020; see also Nyhan & Reifler, 2010; Southwell, Brennen, Paquin, Boudewyns, & Zeng, 2022). Given the strong scientific and medical consensus on vaccines, there is often a clear distinction between what is true versus false (i.e., misinformation).

Although the issue of vaccine hesitancy is not new, there is new urgency in addressing it in the twenty-first century. The 1998 publication of a Lancet article by Andrew Wakefield and colleagues, falsely claiming that the MMR vaccine can cause autism, supercharged vaccine hesitancy despite the article later being retracted (Koslap-Petraco, 2019). False claims about the dangers of vaccines were further shared by prominent celebrities, most notably Jenny McCarthy (Gottlieb, 2015; Largent, 2012). Longstanding news media norms of defining objectivity as providing “two sides” to every story, especially in the United States and many other Western-style democracies, gave this discredited argument unwarranted attention (Clarke, 2008; Dixon & Clarke, 2013).

What makes recent years unique in terms of vaccine hesitancy and the spread of misinformation is the emergence of social media. Social media allows for a democratization of voices, with user-generated content appearing alongside (and often with equal prominence to) official scientific and medical voices. Social media, moreover, can be both insular and porous, allowing diverse views to compete, but also for individuals to often identify and interact primarily with those who share their views (Jones-Jang & Chung, 2022; Schmidt, Zollo, Scala, Betsch, & Quattrociocchi, 2018), phenomenon that has been called the ‘echo chamber.’ Thus misinformation can often live much longer in sheltered corners of social media than it would in the harsher light of public scrutiny. Existing studies have demonstrated that many social media platforms are rife with vaccine misinformation (Suarez-Lledo & Alvarez-Galvez, 2022), which is concerning because misinformation often outperforms accurate information in terms of popularity and reach (Vosoughi, Roy, & Aral, 2018). Perhaps for these reasons, the WHO included vaccine hesitancy as one of the top ten health challenges facing the globe in 2019, even before the emergence of COVID-19 upended public health (WHO, 2019).

COVID-19 created a perfect storm in terms of pre-existing vaccine hesitancy and a media environment that was well-suited to amplify concerns and misinformation about the development of a COVID-19 vaccine that did not have decades or centuries of clear evidence of its specific safety and efficacy. For those who have been engaged in understanding and reducing vaccine hesitancy towards other vaccines, this raises two questions. First, is hesitancy towards the relatively new COVID vaccine – which was developed based on decades of evidence for other vaccines – associated with hesitancy towards other, more established vaccines? Second, is exposure to COVID-19 vaccine misinformation on social media associated with hesitancy towards other vaccines?

We answer these questions in the context of the human papillomavirus (HPV) and hepatitis B (HBV) vaccines because the former has been received with hesitancy by some segments of the public (Saulsberry, Fowler, Nagler, & Gollust, 2019), as was COVID-19. This is due in part, to the politicization of these vaccines. We also include HBV vaccination to determine whether the impact extends to other cancer-related vaccines.

## Methods

A systematic literature review was conducted via the Ovid Medline and APA PsychInfo databases in Fall 2022. We developed a multi-step search strategy (see Table 1) to identify articles that potentially met criteria to answer two research questions: (1) Is COVID-19 vaccine hesitancy, intention, or uptake associated with HPV or HBV vaccine hesitancy, intention, or uptake? (2) Is exposure to COVID-19 vaccine misinformation on social media associated with HPV or HBV vaccine hesitancy, intention, or uptake? Two members of the research team independently completed the multi-step search in each database and achieved consistent results on October 22, 2022. Search steps for the first research question yielded 257 total records (PsychInfo, n=202; Medline, n=55); search steps for the second research question yielded 54 total records (PsychInfo, n=49, Medline: n=5) (see Figure 1).

**Table 1.** Search strategy and number of identified records.

| Search Terms | Medline | PsychInfo |
| --- | --- | --- |
| (1) (misinformation or disinformation or conspiracy theory or rumor or fake news) | 6,248 | 12,908 |
| (2) (social media) or (social network and online) or (social network and digital) or (social network and internet) | 31,155 | 63,944 |
| (3) (COVID) or (SARS-CoV-2) | 304,144 | 32,614 |
| (4) vaccine and (hesitancy or uptake or intention) | 14,105 | 4,856 |
| (5) HPV or HBV | 97,923 | 6,459 |
| Research Question 1: Searches 3, 4, and 5 combined with the Boolean term "and" | 55 | 202 |
| Research Question 2: Searches 1, 2, 3, 4, and 5 combined with the Boolean term "and" | 5 | 49 |

**Figure 1.**
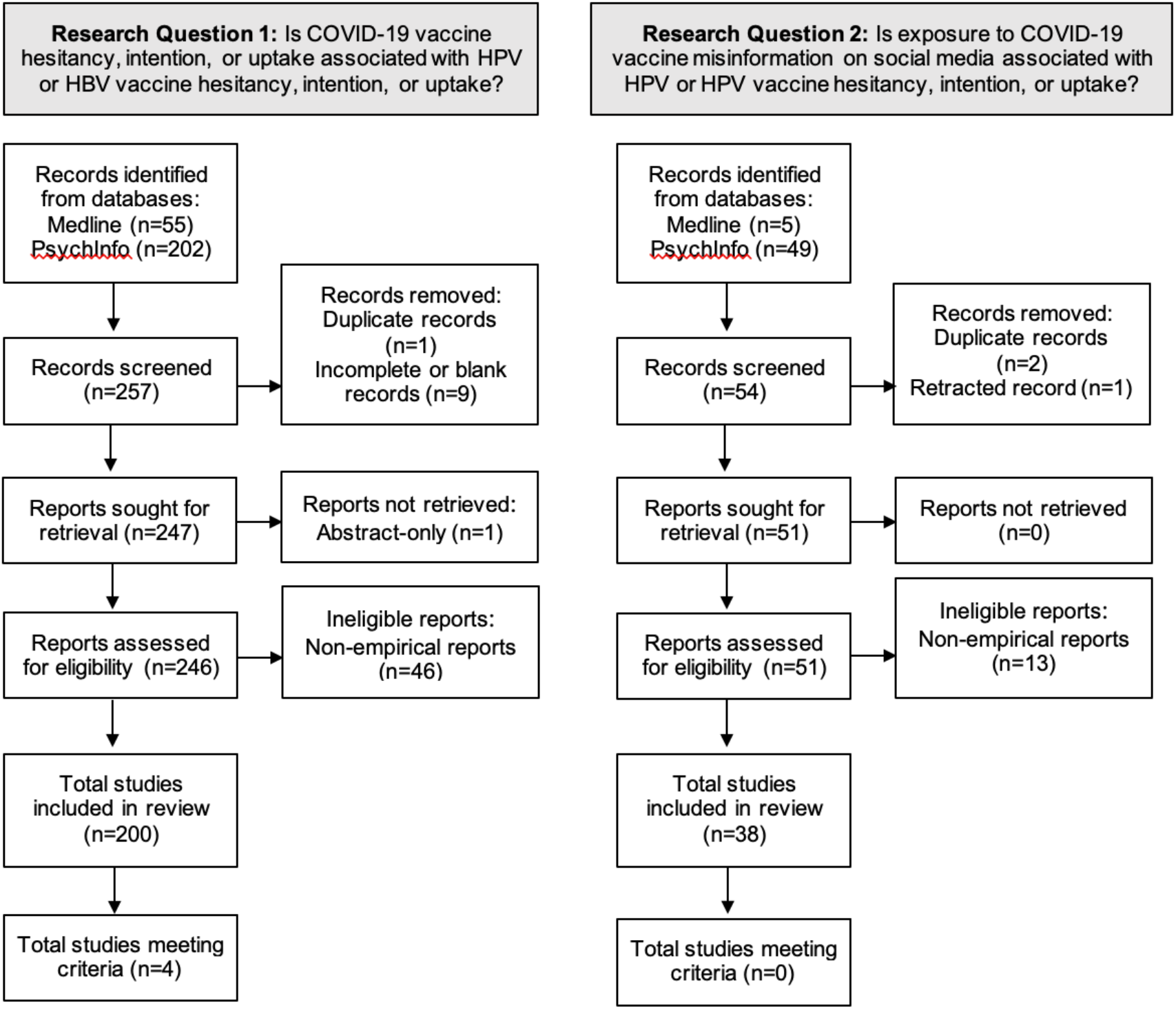
Identification of peer-reviewed articles. *Note: Record* refers to the title or abstract of a report indexed in the Medline or PsychInfo database. *Report* refers to an electronic document providing information about a study, such as a journal article or conference abstract (Page et al., 2021).

For the first research question, 247 reports were eligible for assessment after removing duplicate (n=1) and incomplete (n=9) records. An additional report was unable to be retrieved. To meet criteria for inclusion, a study had to be an empirical study published in an academic journal and (1) measure COVID-19 vaccine hesitancy, intention, or uptake; (2) measure HPV or HBV vaccine hesitancy, intention, or uptake; and (3) test the statistical association between both measures. For the purposes of this review, we defined an empirical study as one that had measures of observable data. Based on these criteria, a further 46 reports were excluded that were commentaries, reviews, opinion pieces, book chapters, simulation modeling, meta-analyses, or animal studies, leaving a total of 200 articles classified as eligible for full review. The reports were divided among seven members of the research team to code independently after first meeting to discuss assessment criteria and reaching consensus on the first 13 articles.

For the second research question, 51 reports were eligible for assessment after removing duplicate records (n=2) and a retracted article (n=1). To meet criteria for inclusion, a study had to be an empirical study published in an academic journal and (1) measure exposure to COVID-19 vaccine misinformation (or disinformation, conspiracy theories, rumors, or fake news) on social media; (2) measure HPV or HBV vaccine hesitancy, intention, or uptake; and (3) test the statistical association between both measures. Based on these criteria, a further 13 reports were excluded, leaving a total of 38 articles classified as eligible for full review. Two members of the research team coded all remaining 38 articles after five members of the research team first met to discuss assessment criteria and reach consensus on 3 articles.

## Results

For our first research question, namely whether COVID-19 vaccine hesitancy, intention, or uptake is empirically associated with HPV or HBV vaccine hesitancy, intention, or uptake, only four of studies of the 200 reports reviewed met our criteria. All these four studies were cross-sectional in nature and examined associations between hesitancy, intention, or uptake for COVID-19 and HPV vaccines, but not the HBV vaccine. Three of these studies found significant associations.

Two of these studies framed their questions in terms of whether COVID-19 constructs impacted HPV constructs, despite being cross-sectional in nature. In January, 2021, Shimuzu and colleagues (2022) conducted a survey of 1,257 Japanese caregivers with daughters aged 12-16 years recruited via a registered research panel. Among other potential determinants of intention to obtain the HPV vaccine for one’s child, the authors measured intention to obtain a COVID-19 vaccine for their child and oneself once the vaccine became available, if side effects were “common” (i.e., not serious). Models were adjusted for demographics, health literacy, media contact, and perceptions and beliefs. Odds of HPV vaccine intention were higher among caregivers who intended to vaccinate their child (OR=4.16, 95% CI: 2.79-6.19) and themselves (OR=1.96, 95% CI: 1.29=3.00) against COVID-19. In March, 2021, Tsui and colleagues (2022) conducted a survey of 357 parents of adolescents aged 9-17 years who were participating in an academic enrichment program for low-income, first-generation, racial or ethnic minority families in Los Angeles, California, United States. Among other potential determinants of HPV vaccine hesitancy, the authors measured intention to obtain a COVID-19 vaccine for their children once it became available. In unadjusted models, the odds of HPV vaccine hesitancy were higher among parents who reported being only somewhat likely (OR=1.58, 95% CI: 0.75– 3.31), not too likely (OR=2.86, 95% CI: 1.16, 7.05), or not likely at all (OR=15.7, 95% CI: 4.45– 55.70) to get their children vaccinated against COVID-19, relative to parents who were very likely to do so. Only the latter association remained significant in models adjusting for covariates, including medical mistrust and exposure to negative information about the HPV vaccine. COVID-19 vaccine intent for children was not associated with HPV vaccine initiation for the youngest child in the family.

Two studies framed their questions in terms of whether HPV constructs impacted COVID-19 constructs but were again cross-sectional in nature. Between November and December, 2020, Berenson and colleagues (2021) surveyed 342 women aged 18-45 years who were recruited from reproductive clinics in South Texas, United States. Adjusting for other factors, women had greater intention to receive a doctor-recommended COVID-19 vaccine if they had previously received the HPV vaccine (OR=2.26, 95% CI: 1.07-4.79). Between March and April, 2021, Phan and colleagues (2022) conducted a survey of 513 caregivers recruited from a pediatric healthcare system in the mid-Atlantic United States. Caregivers were diverse with respect to race, ethnicity, socioeconomic status, and rurality/urbanity. History of child’s receipt of at least one dose of the HPV vaccine if aged 13-20 years was not associated with caregiver intention to vaccinate their children against COVID-19.

Two additional studies did not fully meet criteria for our first research question because they lacked a statistical comparison of the relationship between COVID-19 and HPV (or HBV) vaccine hesitancy, but did explore COVID-19 and HPV constructs in relation to one another. In each study, parents were recruited via an online research panel. Between September and October, 2020, Olagoke and colleagues (2022) conducted a survey of 342 parents of adolescents aged 11-17 years who had never been vaccinated against HPV. To meet eligibility criteria, parents had to identify as Christian and live in the United States. In regression analyses adjusting for sociodemographic variables, perceived vulnerability of one’s child to HPV (β=.32, 95% CI: .21-.44) and perceived response efficacy of the HPV vaccine (β=.41, 95% CI: .28-.53) were independently associated with greater intent to vaccine one’s child against COVID-19, while perceived severity of HPV was not associated with this outcome (β=.16, 95% CI: -.01-.32). All three HPV constructs - perceived vulnerability of one’s child to HPV (β=.37, 95% CI: .25-.48), perceived response efficacy of the HPV vaccine (β=.46, 95% CI: .33-.59), and perceived severity of HPV (β=.21, 95% CI: .05-.38) - were independently associated with parents’ intention to vaccinate oneself against COVID-19. Although intention to vaccinate one’s child against HPV was measured, it was not examined as a predictor of COVID-19 vaccination intentions. In the second study, performed in August 2021, Manganello and colleagues (2022) conducted a survey of 452 parents of children aged 9-14 years living in different communities across the United States. Among parents who would be likely to vaccinate their child against COVID-19, 75% would also be likely to vaccinate their child against HPV. Conversely, among parents who would be likely to vaccinate their child against HPV, 58% would also be likely to vaccinate their child against COVID-19. Although no statistical test was conducted, results suggested that there was greater hesitancy for the COVID-19 vaccine than the HPV vaccine among parents who were accepting of the contrasting vaccine.

Turning to our second research question, our review found that none of the 38 studies reviewed met all three criteria for our second research question, i.e. whether exposure to COVID-19 vaccine misinformation on social media was associated with HPV or HBV vaccine hesitancy, intention, or uptake. Studies identified as potential candidates by our literature search generally described COVID-19 vaccine misinformation on social media, without examining whether exposure to misinformation on social media was associated with HPV or HBV vaccine hesitancy, intention, or uptake among individuals within social media networks.

## Discussion

Our research suggests that there is a dearth of published peer reviewed research studies addressing the question of whether COVID-19 misinformation and vaccine hesitancy spills over to hesitancy towards HPV / HBV vaccines. Only four identified studies examined the association between COVID-19 vaccine hesitancy, intention, or uptake and HPV vaccine hesitancy, intention, or uptake. Moreover, these four studies were all cross-sectional in nature, so that the association between vaccine hesitancy for both vaccines that was uncovered in three out of four studies cannot confidently speak to the directionality of this effect. Since COVID-19 vaccine has been politicized in the United States (Motta, 2021), it is important to further our understanding of how COVID-19 vaccine hesitancy may not only be associated with but also *impact* attitudes towards other vaccines, including those that protect against cancer.

Additionally, there were no peer reviewed studies that explored the association between exposure to COVID-19 *misinformation* on social media and HPV and HBV hesitancy, intention, or uptake. This lack of research is very concerning, especially since so much of the populace relies on social media as a primary information source (Pew, 2022) and social media use is associated with vaccine hesitancy for multiple vaccines (Dunn et al., 2016; Jennings et al., 2021). This area is prime for new empirical research; such findings can inform policy making and regulations to ensure that social media platform policies address health information to promote public health.

## Future Research Directions

There are a number of questions that need to be answered regarding the potential for hesitancy spillover effects across vaccines. Given scholarly and public concerns that COVID vaccine attitudes are impacting uptake for other vaccines (Larson, Lin, & Goble, 2022; Messerly & Mahr, 2022) and alarms about the rising polarization in the US surrounding other vaccines (Francovic, 2021), solid empirical research is necessary to validate a potential link between hesitancy, intention, and especially uptake across vaccines.

Likewise, innovative work is needed to explore the impact of social media exposure to COVID-19 misinformation on HPV and HBV vaccine attitudes. Existing research is often limited to documenting the prevalence of vaccine misinformation on social media (e.g., Suarez-Lledo & Alavarez-Garcia, 2021; Wang, McKee, Torbica, & Stuckler, 2019) or looking at the association between social media use and vaccine attitudes (Dunn et al., 2016; Jennings et al., 2021).

Explicitly linking social media misinformation exposure to individual vaccine beliefs and behaviors will require sophisticated efforts to link online exposure to offline health outcomes. Fortunately, people’s interactions with social media generate vast amounts of detailed behavioral data. Analyzing this data can be extremely useful for developing a fine grained understanding of human beliefs and behaviors (Srivastava, et al 2000), including vaccine hesitancy, intention, and uptake. Future research should document how misinformation, refutation of misinformation, and accurate information about different vaccines propagates through a social network after consecutive sharing (e.g., retweeting) of information. One method is to form a tree structure with the source of information depicted as the root (Figure 2). In this Figure, a follower-followee social network is illustrated where misinformation starts from a source node and propagates through the network when the people exposed to the misinformation share it. This, in essence, forms a propagation tree where the source node is the root of the tree. This same approach can be applied to refutation. As depicted in the Figure, some people are exposed to misinformation only, some are exposed to refutation only, and some are exposed to both, with some people actively spreading misinformation or refutation. Their behavioral actions (to share or not share the true/false information) can vary depending on the order of their exposures, as well as attitude towards misinformation spread. This propagation structure can be utilized to distinguish between true and false information in different domains (Ma et al., 2017, Wu et al., 2015). Looking at individual responses to both misinformation and refutation may allow for better identification of the potential promoters and skeptics of high quality vaccine information.

**Figure 2.**
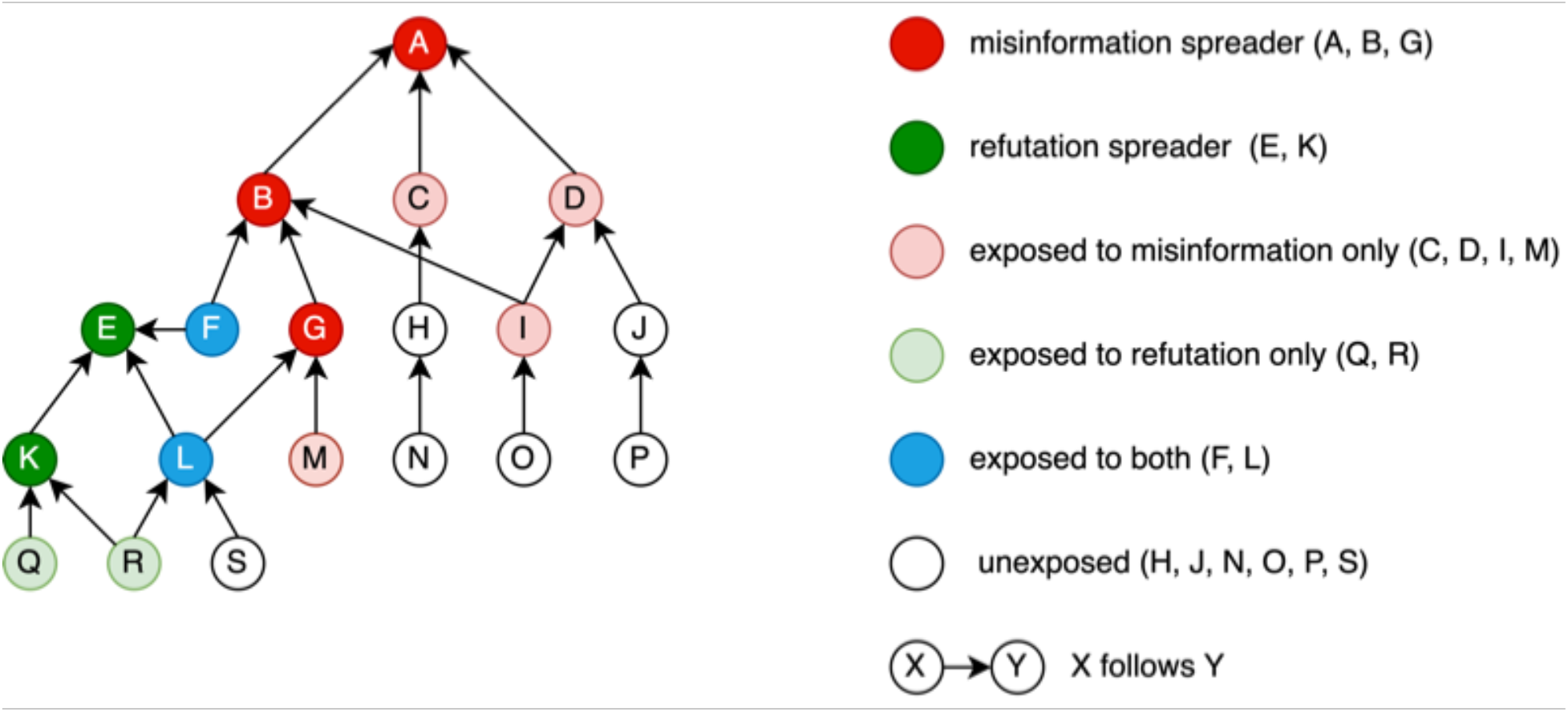
Misinformation and Refutation Propagation Network

To our knowledge, however, investigators have not examined the spread of both vaccine misinformation and refutation in a social network, and whether exposure to both types of information influences vaccine hesitancy, intentions, or uptake. Given the pervasiveness of social media, and the reliance on one’s social media network members to inform decision-making, this type of research is urgently needed. In addition, based on our review, investigators have not attempted to link exposure to COVID-19 vaccine misinformation in a social media network to individual-level HPV or HBV vaccine hesitancy, intention, or uptake.

Other future research directions involve exploring differences in the spread of misinformation on different social media platforms, testing individual and community differences in vulnerability to misinformation spread through social media, and modeling the impact of exposure to misinformation for one vaccine on hesitancy for other vaccines. For example, does the social media platform (e.g., Twitter, Facebook, Instagram, TikTok) impact vaccine hesitancy differently? How does COVID-19 vaccine misinformation spread through social networks? Does COVID-19 misinformation impact its recipients in different ways depending on the source from which they received it? Are some people more susceptible to believing vaccine misinformation and how can we assess their vulnerability? What is the direction of impact in terms of exposure to misinformation about vaccine A impacting hesitancy for vaccine B (i.e., COVID-19 vaccine hesitancy impacting HPV vaccine hesitancy) or vice versa (i.e., HPV vaccine hesitancy impacting COVID-19 vaccine hesitancy)? Putting these pieces together involves developing and testing a comprehensive model that includes social media exposure, vaccine hesitancy, and vaccine uptake for multiple vaccines. Ideally, data would be collected over an extended period of time.

## Conclusion

Tracking people’s interaction with misinformation on social media, and its refutation, provides novel insights into which people are active participants in misinformation spread, and which are active in stopping its spread (Khan et al 2022). This understanding can help in developing strategies to mitigate misinformation’s impact by suppressing the efforts of those who spread misinformation and accelerating the efforts of those who spread the truth. Such prevention strategies are only likely to be effective when there is a foundation of rigorous research to guide efforts.

## Data Availability

There are no data associated with this manuscript.

## Acknowledgements

None

